# Time-of-Day–Dependent Effectiveness of Anti-IgE Therapy in Asthma: Clinical and Experimental Evidence

**DOI:** 10.64898/2026.10.30.26366932

**Authors:** Ayobami Akenroye, Guilin Li, Janice Huss, Carrie Gierasch, Elaine Ostendorf, Scott T. Weiss, Jeffrey A. Haspel

**Author notes:** **CORRESPONDING AUTHOR:** Ayobami Akenroye, MBChB, MPH, PhD, Division of Allergy and Clinical Immunology, Brigham and Women’s Hospital, 60 Fenwood Rd, Boston, MA 02115.

## Abstract

**Rationale:** Asthma is a time-sensitive inflammatory disease whose activity exhibits daily (circadian) rhythms. An outstanding question is whether such temporal variations extend to targeted therapies used for moderate-to-severe asthma.

**Objective:** To examine how time-of-administration relates to the effectiveness of the anti-IgE biologic, omalizumab.

**Methods:** We performed a retrospective cohort study in 153 patients with asthma with timestamped administration records. Time of day was modeled as a continuous variable and dichotomized into morning (8:00-11:59, n=74) versus afternoon (12:00-17:00, n=79) administration. Multivariable logistic regression and Cox time-to-event analyses were used to adjust for key covariates. For biological validation, we treated ovalbumin (OVA)-sensitized mice with anti-IgE antibody at various times of day and measured its impact on inflammatory lung pathology.

**Measurements and Main Results:** Receiving the initial dose of omalizumab in the afternoon was associated with better response, defined as a 50% or more reduction in asthma exacerbations in the year after initiation (Odds ratio, OR: 5.20; 95% CI, 1.94−13.96). For each hour later in the day the initial dose of omalizumab was given, the odds of a response increased by 34% (adjusted OR, 1.34; 1.02–1.77). In OVA-sensitized mice, anti-IgE treatment attenuated inflammatory lung pathology, including OVA-induced airway hyperresponsiveness and type-2 inflammation-related gene expression, in a time-of-day-sensitive manner, with maximum effectiveness when dosed in the latter part of the dark phase, functionally analogous to the afternoon in humans.

**Conclusions:** The timing of anti-IgE treatment for asthma may influence therapeutic success. These findings have implications for optimizing the effectiveness of respiratory biologics.

**At a Glance Commentary:**

- **Scientific knowledge on the subject**: Asthma is a cyclical disease with daily peaks and dips in symptoms and lung function. Frequent nighttime symptoms are a cardinal sign of poor asthma control. Rhythms in lung mechanics coincide with circadian rhythms in autonomic tone, circulating epinephrine, and cortisol. Also, circadian rhythms regulate innate and adaptive immunity, including daily variations in immune cell trafficking, cytokine secretion, pattern recognition receptor signaling, and T-cell activation. Moreover, mast cells and basophils, which express the high-affinity IgE receptor FcεRI, have an intrinsic circadian clock that influences allergic inflammation.
- **What this study adds to the field**: We evaluated if the anti-IgE, omalizumab, varied in effectiveness based on administration time. We found that patients who received their first omalizumab dose in the PM (vs. AM) had better treatment response over the first year of treatment. We validated this finding using circadian-habituated murine models of OVA-sensitized airway inflammation. We found that anti-IgE effectiveness in blunting OVA-induced inflammation and airway responsiveness was dependent on the time of administration. These results suggest that recommending omalizumab dosing in the afternoon could improve omalizumab effectiveness. Our results may have implications for other respiratory biologics.

## INTRODUCTION

Respiratory biologics are monoclonal antibodies that block key mediators of airway inflammation and are increasingly being prescribed for treatment-refractory asthma.[1–3] Currently, there are seven biologics approved for asthma treatment in the US. Of these, the anti-immunoglobulin E (IgE) agent, omalizumab was the first to be approved and is one of the most used worldwide. While randomized trials attest to the efficacy of biologics, post-marketing data reveal that a substantial proportion of patients fail to respond to these agents despite meeting eligibility criteria. While the definition of a positive response to these agents varies, it often includes a reduction in asthma exacerbations by at least half.[4] Based on these criteria, 25-35% of patients fail to respond to asthma biologics after one year of treatment.[5–7] Switching agents and optimizing standard therapies rescue only about 40% of non-responders.[5] Additionally, 30% of patients who previously responded to these biologics cease to benefit after 2 years, resulting in an overall failure rate of up to 40%.[5] This is costly to patients and the healthcare system. Thus, there is a need to improve the clinical performance of biologics.

One emerging approach currently being evaluated for cancer immunotherapies and vaccines is to leverage circadian rhythms in patients’ immune function to enhance treatment success.[8] Circadian rhythms are predictable 24-hour oscillations in biological function thought to emanate from a cell-autonomous molecular “clock”.[8] Circadian rhythms in asthma symptoms have been recognized since antiquity, and frequent nighttime symptoms are a cardinal sign of poor asthma control.[9–11] Rhythms in lung mechanics coincide with circadian rhythms in autonomic tone, circulating epinephrine, and cortisol.[12] Also, circadian rhythms regulate innate and adaptive immunity, including daily variations in immune cell trafficking, cytokine secretion, pattern recognition receptor signaling, and T-cell activation.[13–15] Moreover, mast cells and basophils, which express the high-affinity IgE receptor FcεRI, have an intrinsic circadian clock that influences allergic inflammation.[16, 17]

Clinically, circadian influences can manifest as a therapy exhibiting differential effectiveness based on the time-of-day it is administered. For example, the potency of inhaled corticosteroids in asthma varies by time-of-day, with late-afternoon or evening dosing favored.[10, 18, 19] Here, we postulated that the time-of-day could influence the performance of omalizumab. Thus, we combined observational human data and an experimental mouse model of type-2 lung inflammation to examine whether anti-IgE therapy for asthma varies in effectiveness across time of day.

## METHODS

### A. Anti-IgE (omalizumab) retrospective cohort study

#### Data Source and Population

We used data from the Effectiveness of Respiratory biologics in Asthma (ERA) cohort, a growing retrospective cohort of monoclonal antibody-naïve patients with moderate-to-severe asthma seen by an allergist or pulmonologist in the Mass General Brigham Healthcare system (MGB), a large health system in Northeast America.[20] We limited the data to patients who initiated omalizumab between January 1, 2016, and December 31, 2024, and who had timestamped administration records available. We excluded any patient with a concomitant diagnosis of chronic urticaria, another leading indication for omalizumab, to ensure that asthma was the predominant reason for omalizumab prescription. We also excluded patients who did not receive their first injection within the first year of the prescription, as these patients may differ from those who received their injection within a year of therapy (**Fig. 1A**).[21] Subsequently, given that 98% of injections were administered in the block between 8:00 and 17:00, we limited to patients who received their dose within this timeframe (**Fig. 1B**).

**Figure 1.**
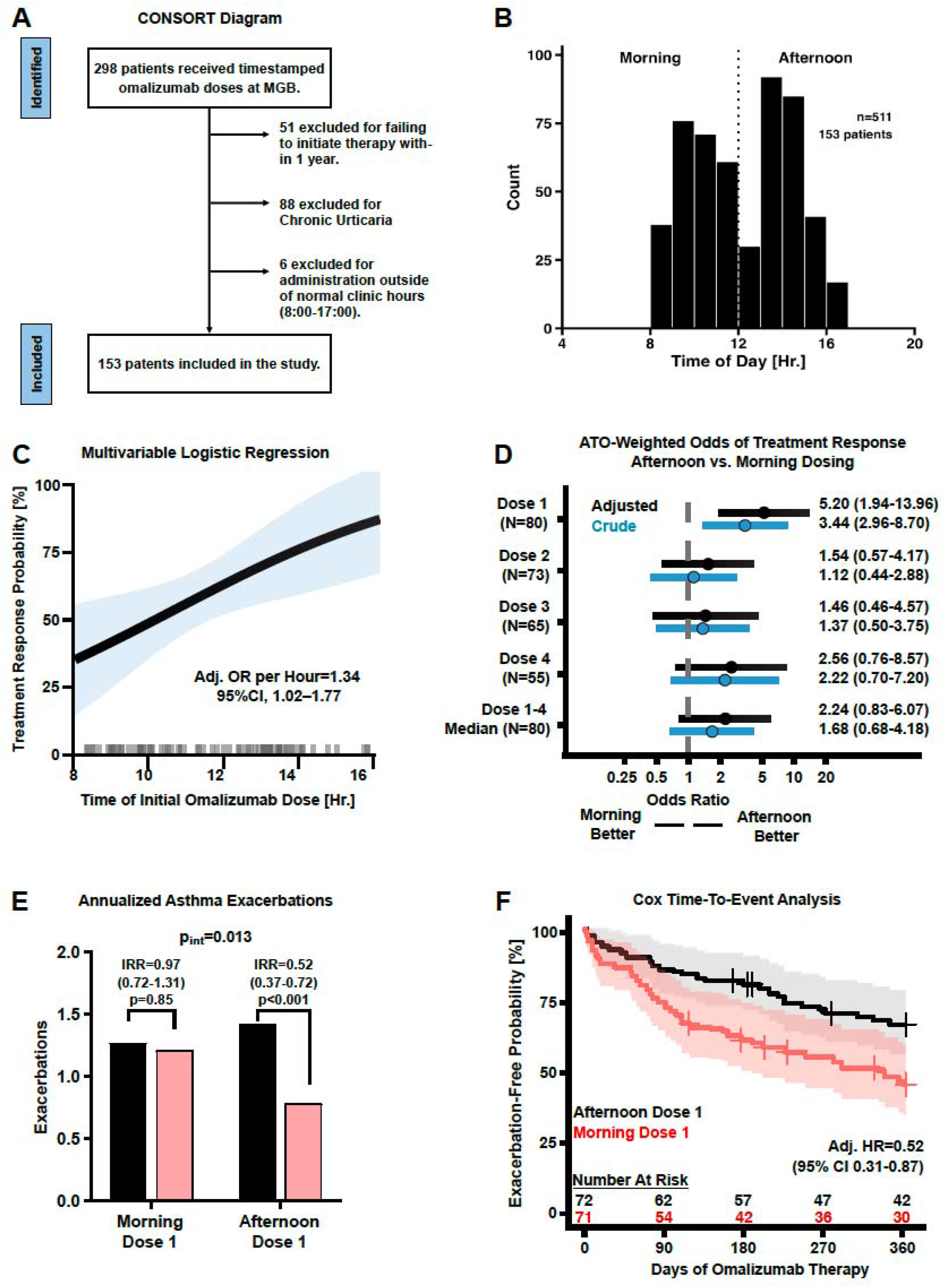
Association between omalizumab clinical effectiveness and time of day. (A) CONSORT inclusion/exclusion diagram for our retrospective cohort. (B) Histogram showing the distribution of omalizumab administration times for the first four doses (n=511 doses). (C) Multivariable logistic regression showing the relationship between the time of administration of the first omalizumab dose and treatment response, defined as a 50% or greater reduction in asthma exacerbations during the first year of therapy. In this analysis, time of day is treated as a continuous variable. The average hourly increase in the adjusted odds ratio (OR) of a treatment response, with 95% CI, is shown in the panel. 8:00-16:00 shown for visualization purpose; model included all individuals who received doses between 8:00 and 17:00. (D) Propensity (ATO)-weighted analysis of omalizumab treatment response based on the time of day of doses 1, 2, 3, 4, or the median time of day for doses 1-4. Blue bars, unadjusted OR; black bars, adjusted OR. (E) Change in annualized asthma exacerbations after omalizumab treatment, binned by morning (N=74) vs. afternoon initiation of omalizumab (N=79). Incidence rate ratios (95% CI) and a p-value for the unadjusted interaction between treatment response and time-of-day is shown. (F) Cox regression analysis of the asthma exacerbation-free probability over time, based on the initial omalizumab dose given in the evening (black line) versus the morning (red line). Shaded areas represent 95% CIs. The exacerbation-free OR associated with afternoon omalizumab initiation, with 95% CI, is provided.

#### Exposure and Outcomes

The primary exposure was the timing of the first four doses of therapy. This was modeled first as a continuous variable using the actual time of administration and as a binary variable: morning (8:00-11:59) and afternoon dosing (12:00-17:00). We focused on the initial four doses given initial evidence from immunotherapy in cancers that the chronotherapeutic effect is particularly tied to the timing of the first dose,[22] pharmacokinetic modeling showing that most therapies reach steady state after about 4-5 elimination half-lives and that patients on omalizumab may reach steady state by the fourth to sixth dose,[23, 24] and to maximize data availability given that the first three doses of omalizumab are often administered in the clinic.

We defined clinical response as a 50% reduction in asthma-related exacerbations over 12 months of therapy. Exacerbations were defined as an acute loss of asthma control necessitating an asthma-related emergency room visit, admission, or a course of oral corticosteroids for three or more days.[25] In negative binomial models, we used exacerbation count as the outcome, without defining a threshold for response; and in time-to-event analyses, the outcome was the number of days from the first omalizumab prescription to the first post-prescription exacerbation event.

#### Statistical Analyses

Logistic regression models were used to evaluate the association between dosing time and response limiting to patients who had one or more exacerbations in the year prior to baseline. We used negative binomial models to compare the change from the year before to the year after first omalizumab prescription (incidence rate ratios, IRR) with an interaction term to evaluate whether this differed by afternoon vs. morning dosing. Finally, we evaluated the association between dosing time and time to first exacerbation using a Cox regression model. As we have previously done, and as recommended to enhance equipoise between comparator populations,[25–28] we used propensity score overlap weighted analyses to balance measured covariates between the two dosing groups, thus focusing on patients with a substantial probability of receiving treatment at either timeframe. The weighting model included prespecified covariates potentially associated with both the timing of administration and the outcome. All models included age, sex, body mass index (BMI), smoking status, insurance type, race-ethnicity, baseline exacerbation rate, baseline eosinophil count, comorbid chronic obstructive pulmonary disease (COPD), concomitant chronic rhinosinusitis or nasal polyps (CRSwNP), and Charlson Comorbidity Index (CCI). Where administration time was modeled as a continuous variable, we used multivariable adjustment to account for these confounders. Subsequently, we estimated the predicted probability of response for each one-hour increment in dosing time. Likewise, we estimated the marginal probability of response if all patients received their doses at each hour of the 9-hour block of time, the predicted probability of response for afternoon vs morning dosing, and the probability of remaining exacerbation-free at 1 year.

#### Stratified and Sensitivity Analyses

We performed stratified analyses by treatment site (Brigham and Women’s Hospital vs. other MGB hospitals), and dosing interval (biweekly vs. monthly). In sensitivity analyses, we evaluated how results and model performance were affected by: 1) using inverse probability treatment weighting (IPTW) instead of propensity score overlap weighting; 2) using splines and modeling time as a circular variable; 3) adding baseline IgE level as a model covariate; 4) adding season of administration as a model covariate,[29] and 5) by dose timing consistency according to patterns across multiple doses (afternoon/mixed vs. consistently morning administration, among patients with ≥2 doses). Finally, we evaluated the impact of multiple imputation by chained equations for eosinophil count and IgE for patients missing these variables on our results. We compared the performance of nested models (with vs. without adjustment) using the Akaike Information Criterion (AIC) and the Likelihood Ratio Test (LRT).[30]

All above analyses were performed using R version 4.5.1.[31] ChatGPT (OpenAI) was used to assist with debugging R code. All AI-assisted code was reviewed, verified, and executed by the authors, who independently evaluated the analytical methods and results. The authors take full responsibility for the accuracy, integrity, and interpretation of the analyses. The MGB Institutional Review Board (IRB) approved this clinical study (#2023P003017). See Appendix (Section 1) for additional methods.

### B. Murine Studies

To biologically validate our retrospective observations, we modeled allergic asthma in female BALB/C mice using an adjuvant-free Ovalbumin (OVA) sensitization protocol (**Fig 2, A**).[32]

**Figure 2.**
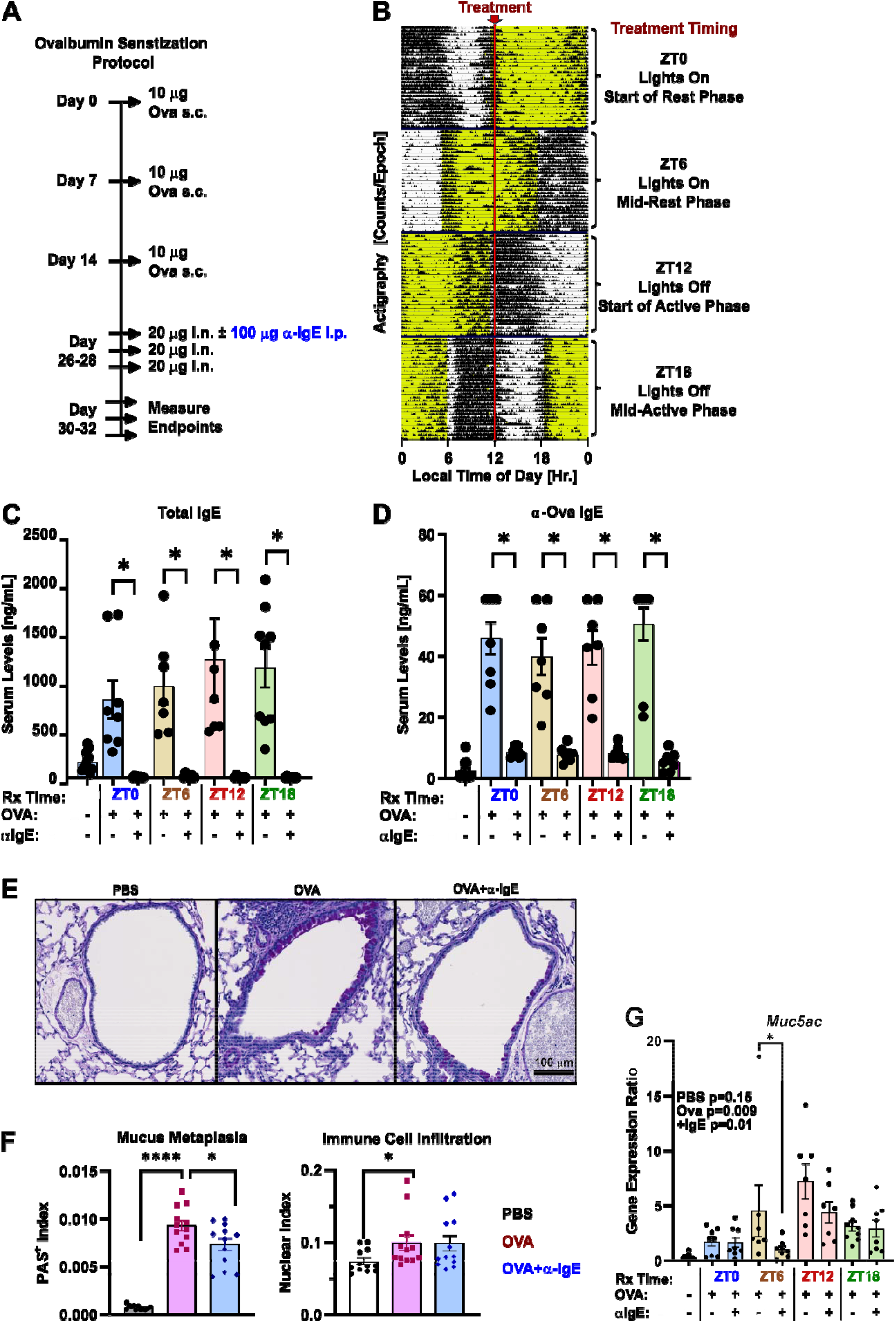
Model for examining time of day variation in anti-IgE treatment for allergic asthma. (A) OVA sensitization and exposure timeline. A single dose of anti-IgE (blue lettering) or sham (PBS) was given as an intraperitoneal (i.p.) injection 20 minutes before the first intranasal OVA exposure. (B) Representative actigraphy in group-housed mice (n=4/cage) throughout the OVA sensitization protocol (panel A), passively measured as counts per 10-minute epoch (black bars). Lighted intervals of the day for each of four treatment groups are highlighted in yellow. Each row represents 24 hours of observation, with 30 days of telemetry provided per group. The time of experimental treatments (Ova or a-IgE) is denoted by a vertical red line and expressed in units of zeitgeber time to the right of each treatment group (ZT0=lights on, ZT12=lights off). (C, D) Serum total IgE (C) and Ova-specific IgE (D) as a function of treatment time of day (mean ± SE, n=3-10) pooled from two independent experiments. *p<0.05, two-tailed Student’s t-Test of log-transformed data. (E) Representative PAS-stained micrographs of lungs from Sham (PBS), Ovalbumin (Ova), and Ova+a-IgE treated mice. Scale Bar=100 mm. (F) Morphometric quantification of PAS^+^ (left panel) and Hematoxylin^+^ (nuclear) density (right panel) reflecting the extent of mucus cell metaplasia and immune cell infiltration, respectively (mean ± SE, n=12 drawn equally across all treatment times of day). Black symbols, PBS-treated; red symbols, Ova-treated, blue symbols, Ova+a-IgE treated. *p<0.05, ****p<0.001, Student’s 2-tailed t-test on log-transformed data. (G) *Muc5ac* lung gene expression based on the time of day of OVA sensitization and a-IgE treatment (mean ± SE, n=9-12). Note that Sham (PBS) mice have been pooled across ZT times for visual clarity. One-way ANOVA p-values for log-transformed data are provided in the legend. *p<0.05, Student’s 2-tailed t-test on log-transformed data. Data are pooled from two independent experiments.

#### Mice

Six-week-old female BALB/C mice (20-25 g) were obtained from Jackson Laboratory. Mice were housed in shoebox-sized cages with food available *ad libitum*. To adjust the circadian phases of mice, they were acclimated in custom light-tight enclosures (Gorilla Grow Tent) for two weeks with the light cycles controlled by a programmable timer as described.[33] Mouse actigraphy was recorded in individually housed mice using passive IR monitors, and data were analyzed using ClockLab software (Actimetrics). The Washington University School of Medicine Institutional Review Board (#24-0180) approved all experiments.

#### Ovalbumin (OVA) sensitization

We employed an adjuvant-free OVA sensitization protocol to experimentally model asthmatic lung pathology as described.[32] To add anti-IgE (α-IgE) therapy and time of day to this model, the mice were habituated to one of four staggered light-dark cycles before OVA sensitization.[33] This had the effect of shifting the circadian cycles of the mice, such that treatments given at 12:00 local time represented different biological times of day, depending on which light cycle the mice were housed in (**Fig 2, B**). OVA sensitization commenced after mice acclimatized for two weeks in custom light cycles to simulate different exposure times of day (**Fig 2, A**). All treatments were administered at 12:00 local time under dim red lighting to avoid blue-light exposure that could shift the mice’s circadian phase. In circadian rhythm research, internal biological time is represented in “Zeitgeber Time” (ZT) units, where ZT0 is lights-on, and ZT12 is lights-off.[34] Mice were anesthetized with 2% isoflurane and received subcutaneous injections of OVA (10 mg/100 mL sterile PBS) on days 0, 7, and 14 of the protocol. On Days 26, 27, and 28, they were anesthetized and intranasally dosed with 20 mg OVA in 30 mL of sterile PBS. To simulate omalizumab therapy, mice received a single (100 mg/100 mL) dose of (α-IgE) monoclonal antibody (clone R35-72, BD Biosciences) at ZT0, ZT6, ZT12, or ZT18, just before the first OVA inhalational exposure (**Fig 2, A**), thereby simulating dose 1 of omalizumab (**Fig 1, D**). Sham (PBS) injected mice served as controls.

#### Respiratory system mechanics measurement

Respiratory system mechanics were measured using a Buxco Elan RC mouse volume-controlled ventilator as described.[35]

#### Quantitative PCR (qPCR)

RNA extraction and qPCR were performed using standard protocols.[36] Primers used for qPCR are listed in **Table S1**.

#### Histology and Immunostaining

Lungs were fixed by inflation with 10% buffered formalin at 20 cmH2O, paraffin-embedded, and stained with Periodic Acid Schiff Stain as described.[35] Slides were scanned using a Hamamatsu Nanozoomer at the Alafi Neuroimaging lab at Washington University. PAS-positive and hematoxylin-positive pixels were counted using Aperio ScanScope Software as described.[37]

#### Statistics

For all mouse-derived data, one-way ANOVA and Student’s 2-tailed t tests were performed on log-transformed data using Microsoft Excel or GraphPad Prism software. Blinding was employed for all experiments. For all endpoints, data were pooled from at least two independent experiments. See Appendix (Section 2) for additional methods.

## RESULTS

### Omalizumab’s effectiveness correlates with the time of day at which treatment is initiated

We identified 298 patients in ERA receiving omalizumab for asthma for whom timestamps were recorded for one or more of the first 4 doses. Of these, 51 patients were excluded for not initiating treatment within a year of prescription, 88 patients were excluded because of concomitant chronic urticaria, and 6 patients administered outside of normal outpatient clinic hours (8:00 to 17:00) (**Fig 1A**). Our final cohort comprised 153 patients, with 48.4% receiving their first dose during the morning clinic session (**Table 1**, **Fig 1B**). In terms of dosing frequency, most patients received monthly doses of omalizumab, while 33 of 153 (21.6%) received biweekly doses (**Fig S1**). Patients who received their first dose of omalizumab in the afternoon vs. the morning had similar demographics, clinical characteristics, laboratory values (eosinophil count, IgE), dosing frequencies, and treatment sites (**Table 1**).

**Table 1:** Baseline characteristics of patients according to timing of their first omalizumab administration.

| Characteristic | Overall | Morning | Afternoon | P value |
|---|---|---|---|---|
| N | 153 | 74 | 79 |  |
| Age in years, mean (SD) | 42.4 (18.9) | 42.3 (19.2) | 42.6 (18.7) | 0.92 |
| Female sex, n (%) | 111 (72.5%) | 54 (73.0%) | 57 (72.2%) | 1.00 |
| White race, n (%) | 117 (76.5%) | 55 (74.3%) | 62 (78.5%) | 0.55 |
| Hispanic ethnicity, n (%) | 8 (5.2%) | 4 (5.4%) | 4 (5.1%) | 1.00 |
| Race/ethnicity, n (%) |  |  |  | 0.86 |
| All Other | 47 (30.7%) | 22 (29.7%) | 25 (31.6%) |  |
| White non-Hispanic | 106 (69.3%) | 52 (70.3%) | 54 (68.4%) |  |
| Private insurance, n (%) | 117 (76.5%) | 58 (78.4%) | 59 (74.7%) | 0.70 |
| Body mass index (BMI) in kg/m<sup>2</sup>, mean (SD) | 28.3 (6.7) | 28.2 (7.1) | 28.4 (6.5) | 0.70 |
| Current/Former smoker, n (%) | 12 (7.8%) | 7 (9.5%) | 5 (6.3%) | 0.80 |
| Charlson Comorbidity Index, mean (SD) | 1.4 (1.5) | 1.4 (1.1) | 1.4 (1.8) | 0.41 |
| Comorbid conditions, n (%) |  |  |  |  |
| COPD | 12 (7.8%) | 7 (9.5%) | 5 (6.3%) | 0.47 |
| Chronic sinusitis | 14 (9.2%) | 4 (5.4%) | 10 (12.7%) | 0.12 |
| Nasal polyposis | 6 (3.9%) | 5 (6.8%) | 1 (1.3%) | 0.11 |
| Chronic sinusitis/nasal polyposis | 19 (12.4%) | 8 (10.8%) | 11 (13.9%) | 0.56 |
| Exacerbations in the year before biologic prescription, mean (SD) | 1.3 (1.7) | 1.3 (1.6) | 1.3 (1.8) | 0.97 |
| Exacerbation counts in year before prescription (with or without admission), n (%) |  |  |  | 0.93 |
| 0 | 70 (45.8%) | 34 (45.9%) | 36 (45.6%) |  |
| 1 | 35 (22.9%) | 16 (21.6%) | 19 (24.1%) |  |
| ≥2 | 48 (31.4%) | 24 (32.4%) | 24 (30.4%) |  |
| Asthma admission counts in year before prescription, n (%) |  |  |  | 0.56 |
| 0 | 140 (91.5%) | 69 (93.2%) | 71 (89.9%) |  |
| 1 | 10 (6.5%) | 3 (4.1%) | 7 (8.9%) |  |
| ≥2 | 3 (2.0%) | 2 (2.8%) | 1 (1.3%) |  |
| <sup>a</sup>Blood eosinophil count in cells/mcl |  |  |  | 0.15 |
| Mean (SD) | 223.7 (245.3) | 254.2 (260.6) | 193.7 (227.1) |  |
| Median (Q1, Q3) | 150 (60, 280) | 190 (70, 340) | 125 (60, 230) |  |
| <sup>a</sup>Total IgE in IU/ml |  |  |  | 0.91 |
| Mean (SD) | 343.0 (703.8) | 456.3 (973.2) | 260.2 (347.6) |  |
| Median (Q1, Q3) | 152 (52, 346) | 146 (44, 442) | 155 (64, 294) |  |
| Dosing interval, n (%) |  |  |  |  |
| Monthly | 104 (68.0%) | 51 (68.9%) | 53 (67.1%) | 0.97 |
| Biweekly | 33 (21.6%) | 15 (20.3%) | 18 (22.8%) |  |
| <sup>b</sup>Unclassified | 16 (10.4%) | 8 (10.8%) | 8 (10.1%) |  |
| Institution, n (%) |  |  |  | 0.86 |
| BWH | 87 (56.9%) | 43 (58.1%) | 44 (55.7%) |  |
| MGH | 45 (29.4%) | 22 (29.7%) | 23 (29.1%) |  |
| All other | 21 (13.7%) | 9 (12.2%) | 12 (15.2%) |  |
| Season, n (%) |  |  |  | 0.50 |
| Winter | 50 (32.7%) | 22 (29.7%) | 28 (35.4%) |  |
| Spring | 41 (26.8%) | 24 (32.4%) | 17 (21.5%) |  |
| Summer | 27 (17.6%) | 12 (16.2%) | 15 (19.0%) |  |
| Fall | 35 (22.9%) | 16 (21.6%) | 19 (24.1%) |  |

BWH, Brigham and Women’s Hospital; COPD, chronic obstructive pulmonary disease; IgE, immunoglobulin E; MGH, Mass General Hospital; Q1, first quartile; Q3, third quartile; SD, standard deviation.
<sup>a</sup>10 patients (3 in morning group and 7 in afternoon group were missing baseline eosinophil counts. 28 patients (15 in morning group and 13 in afternoon group were missing baseline IgE level.
<sup>b</sup>Received a single dose.

The time-of-day of the first omalizumab dose was associated with treatment response in the 12 months following treatment initiation. For each hour later in the day the initial dose was given, the odds of a treatment response increased by 34% (OR: 1.34; 95% CI, 1.02–1.77) (**Fig 1C**; **Table 2**). Modeling time of day as a dichotomous rather than a continuous variable yielded similar results, with afternoon initiation associated with greater response (**Table 3**). In propensity score overlap-weighted analyses with good covariate balance (**Fig S2**), afternoon initiation of omalizumab therapy was again associated with better response compared to morning dosing (adjusted OR 5.20; 95% CI 1.94-13.96) (**Fig 1D**), with a predicted probability of response over 12 months of 76.3% vs. 38.2% (**Fig S3, A**). Notably, we did not observe a significant association between the time of day of doses 2-4 and response (**Fig 1D**). Comparing the incidence rate ratio (IRR) of asthma exacerbations before omalizumab prescription to the year after, those who received their first dose in the morning did not experience significant reductions in exacerbation rates (1.26 pre to 1.22 post, IRR, 0.97; 95% CI: 0.72-1.31) while those in the afternoon group had a significant reduction in exacerbations (1.33 to 0.69, IRR 0.52, 95% CI: 0.37-0.72; p for interaction= 0.013) (**Fig 1E**). For doses 2–4, the IRRs did not differ substantially between afternoon and morning administration (**Table S2**).

**Table 2:** Association between omalizumab administration time (as a continuous variable) and response in 12 months of follow up.

| Predictor | N | Adjusted Odds ratio (OR)<br>Afternoon vs morning | 95% Confidence interval: Lower | 95% Confidence interval: Upper | p-value |
|---|---|---|---|---|---|
| **Dose 1** | **80** | **1.34** | **1.02** | **1.77** | **0.036** |
| Dose 2 | 73 | 1.23 | 0.97 | 1.55 | 0.093 |
| Dose 3 | 65 | 1.24 | 0.98 | 1.57 | 0.075 |
| Dose 4 | 55 | 1.25 | 0.96 | 1.61 | 0.092 |
| **Median first 4** | **80** | **1.46** | **1.11** | **1.92** | **0.007** |

**Table 3:** Inverse Probability Treatment Weighted (IPTW) estimates of association between dosing time (afternoon vs. morning) and response. (NB: negative binomial).

| **Timing** | **Logistic: Odds ratio, OR (95% CI)** | **Cox: Hazard ratio, HR (95% CI)** | **NB Ratio of Incidence Rate Ratio, IRRs (95% CI)** |
|---|---|---|---|
| **Dose 1** | **5.20 (1.97-13.72)** | **0.54 (0.32-0.92)** | 0.56 (0.25-1.26) |
| Dose 2 | 1.59 (0.59–4.34) | 0.94 (0.52–1.71) | 1.72 (0.72–4.07) |
| Dose 3 | 1.91 (0.59–6.13) | 0.90 (0.52–1.57) | 1.41 (0.77–2.57) |
| Dose 4 | 3.09 (0.91–10.47) | **0.44 (0.23–0.85)** | 0.51 (0.25–1.03) |
| Median first 4 | 2.17 (0.80–5.89) | 0.77 (0.41–1.43) | 1.17 (0.43–3.19) |

Similar results were observed in time-to-event analyses (**Fig 1F**). Afternoon omalizumab initiation was associated with a lower hazard of asthma exacerbations, with the afternoon-initiation group more likely to be exacerbation-free at 1 year (HR 0.52, 95% CI 0.31–0.87). There was no significant association between response and timing of administration with doses 2 and 3. However, afternoon administration of dose 4 was also associated with a lower hazard of asthma exacerbations over one year (HR 0.46, 95% CI 0.24–0.89) (**Figure S4, A-C**).

In sensitivity analyses, inverse probability treatment weighted (IPTW) analyses were consistent with the propensity score overlap-weighted analyses, with afternoon administration of dose 1 associated with better response than morning administration (OR 5.20, 95% CI: 1.97-13.72, **Table 3**). Patients who received their first two or more doses of omalizumab in the afternoon had higher odds of a clinical response than patient who received exclusively morning doses (OR: 4.51, 95% CI 1.45-14.01). The association between timing and response was similar among patients receiving predominantly monthly versus biweekly omalizumab (OR 5.19 (1.57–17.15) vs 1.14 (0.17–7.76); P for interaction = 0.19), though only 19 (26%) of the 73 patients with at least one baseline exacerbation and complete covariates were in the biweekly group. Addition of serum IgE as a model covariate did not significantly improve model fit for treatment response, exacerbation rate, or time to first exacerbation (**Table S3**). Moreover, results were consistent following imputation of missing IgE and/or eosinophil values (**Table S4**). Likewise, results were robust to the addition of season of administration (**Table S5**). Results were similar across treatment initiation sites (**Table S6; Figs S5-S7**). Finally, there was no evidence that spline or circular time specifications improved model fit (**Table S7**). Altogether, our retrospective analysis found that omalizumab effectiveness in asthma tracked with the time-of-day therapy was initiated, with afternoons associated with treatment success.

### Anti-IgE treatment against asthmatic lung pathology in mice is also time-of-day sensitive

We proceeded with biologic validation. OVA sensitization induced high total and OVA-specific serum IgE levels, which were suppressed by α-IgE treatment, independent of the time of day it was administered (**Figs 2, C** and **D**). Histologically, OVA induced mucus cell metaplasia and peri-bronchial immune cell infiltration across all treatment times of day, and this was mitigated by α-IgE (**Figs 2, E and F**). However, the induction of inflammatory mucus (*Muc5ac*) by OVA varied significantly in magnitude based on the time of day at which mice were exposed to the allergen (**Fig 2, G**). Moreover, the ability of anti-IgE treatment to attenuate this induction varied with time of day, being most potent with ZT6 dosing (**Fig 2, G**).

We next assessed airway hyperresponsiveness (AHR) to methacholine (**Fig 3**), which, in murine asthma models, reflects the cumulative mechanical impact of airway smooth muscle hyperreactivity, lung edema from type-2 inflammation, and mucus plugging.[38] OVA induced AHR to equal extents regardless of sensitization timing. (**Fig 3, A-D**, red versus black lines), However, α-IgE was only effective at blunting AHR when administered to OVA-sensitized mice at ZT18 (**Fig 3, D**).

**Figure 3:**
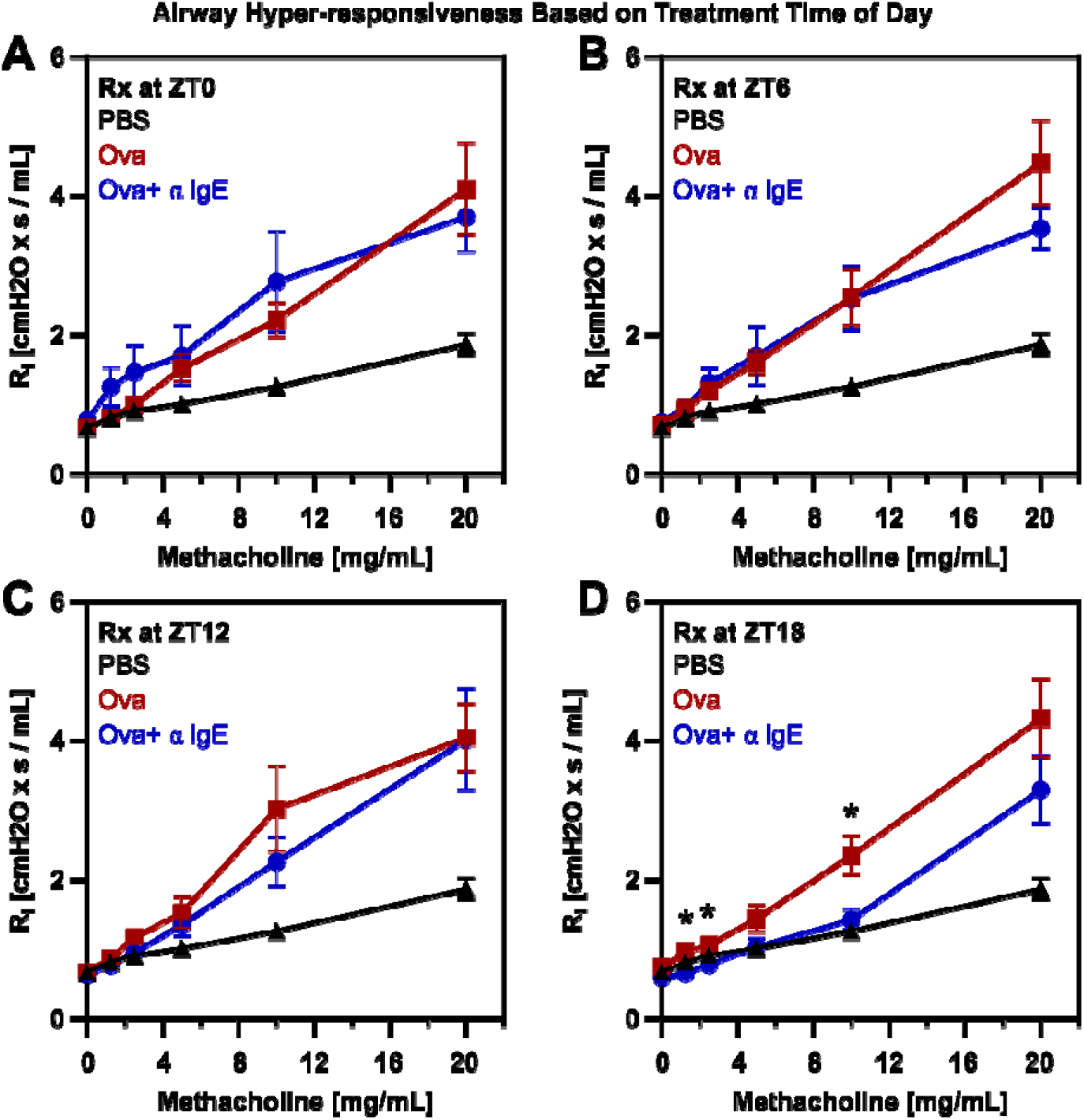
Anti-IgE treatment mitigates airway hyperresponsiveness (AHR) in a time-of-day-dependent manner in mice. (A-B) Plots of dynamic inspiratory resistance versus concentration of nebulized methacholine in Ova-sensitized mice AHR as a function of OVA and a-IgE treatment ZT time (mean ± SE, n=6-11). An increase in the slope of the airway resistance (R_I_) x methacholine dose-response indicates airway hyperresponsiveness. Black, sham (PBS) treated mice; Red, OVA treated mice; Blue, Ova+-IgE. *p<0.05 OVA vs. OVA + a-IgE (Student’s 2-tailed t-test of log-transformed data). Data are pooled from two independent experiments.

Turning to mediators of type-2 inflammation (**Fig 4**), the induction of inflammatory gene expression by OVA showed significant time-of-day variation for *IL13*, *Arg1*, and *Eotaxin* (**Fig 4, A-C**), but not for *IL-5* or *IL-33* (**Fig 4, D and E**). In all cases, α-IgE treatment did not uniformly mitigate the induction of these genes throughout the day; instead, it was effective at specific treatment times. Using a panel of inflammatory markers, we observed that α-IgE treatment at ZT18 produced the most consistent suppression of type-2 inflammation at the gene expression level, followed by dosing at ZT12 (**Fig. 4, F**). Interestingly, a-IgE treatment also upregulated certain inflammatory mediators in a time-of-day-specific manner; for example, ZT18 dosing uniquely upregulated *IL6* expression (**Fig. 4, F**). In contrast, α-IgE treatment at ZT6 upregulated *Ifng* and *Cd4* expression, suggesting greater infiltration of activated T cells. Altogether, our data demonstrated a temporal gradient in OVA’s ability to induce type-2 lung pathology and, separately, the ability of IgE antagonism to suppress that pathology. The cumulative result was that anti-IgE treatment was effective in mitigating allergic lung pathology when administered at some times of day (ZT18) but less so at others (ZT0).

**Figure 4:**
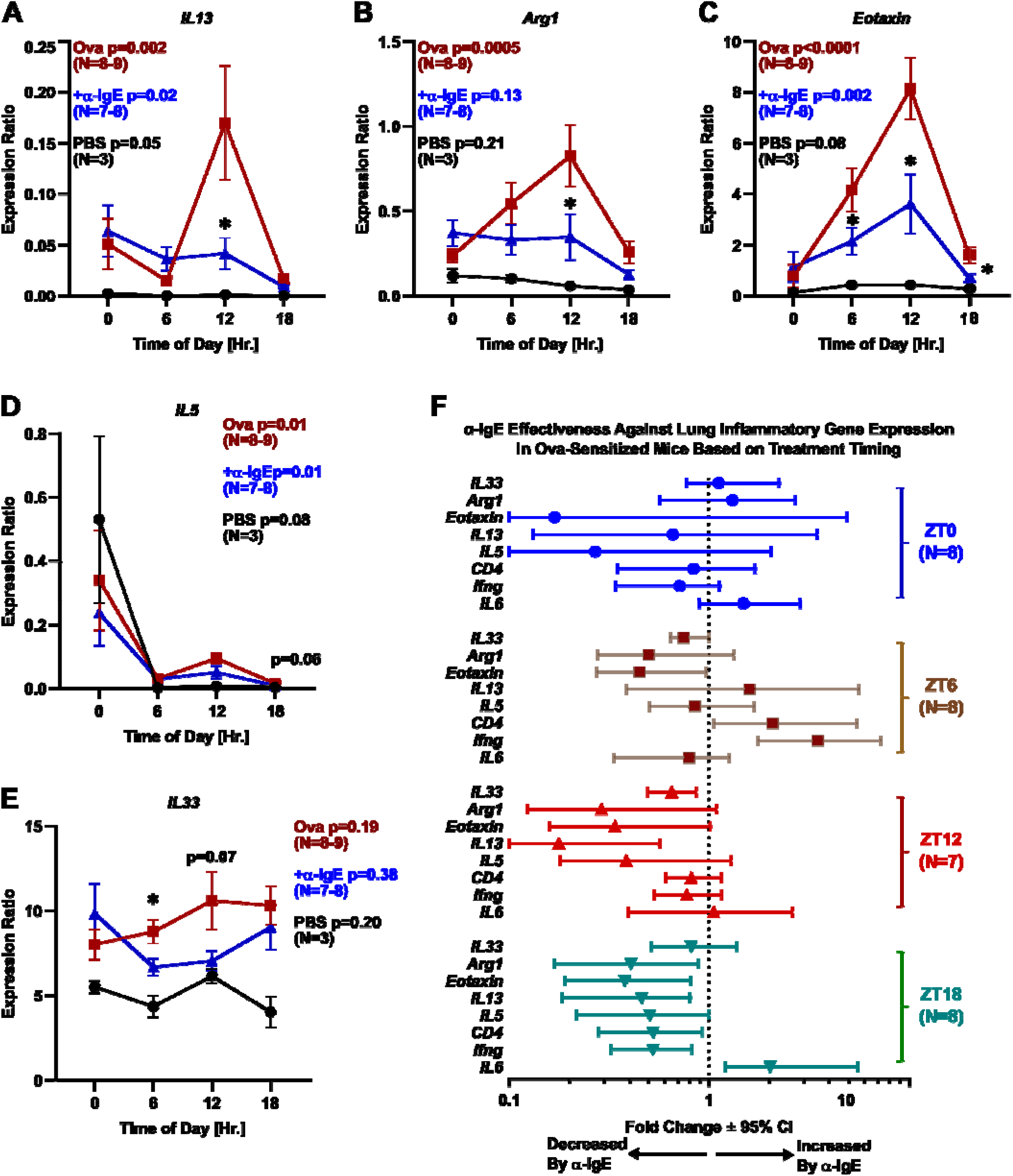
Time of day influences the effectiveness of IgE antagonism against type-2 lung inflammation in mice. (A-E) Lung gene expression of various type-2 inflammatory mediators as a function of treatment (PBS, black symbols; OVA sensitization, red symbols; OVA + a-IgE, blue symbols) and administration time of day expressed in ZT units (mean ± SE, n=3-8). One-way ANOVA p-values on log-transformed data are provided in the legends. *p<0.05 OVA vs. OVA + a-IgE (Student’s 2-tailed t-test of log-transformed data). (A) *IL13*. (B) *Arg1*. (C) *Eotaxin*. (D) IL5. (E) *IL33*. (F) Relative change in gene expression of various inflammatory markers, comparing Ova-sensitized mice treated at different times of day with a-IgE to OVA-sensitized mice alone (n=7-8). Symbols represent the mean ± 95% CI plotted on a log scale. Deviations to the left of the dotted line indicate relative suppression by a-IgE treatment, while deviations to the right represent relative induction by a-IgE treatment. Data are pooled from two independent experiments.

## DISCUSSION

To our knowledge, this is the first report that a biologic approved for asthma can vary in effectiveness based on the time of day it is administered. We focused on the anti-IgE biologic, omalizumab, in this study because it is more frequently initiated in the clinic setting than other agents, and therefore timestamps were more readily available for time-of-day comparisons. Our findings have direct implications for omalizumab indications beyond asthma, such as IgE-mediated food allergies, as well as for other respiratory biologics that target alternative mediators of airway inflammation. Specifically, we found anti-IgE effectiveness was higher when initial dosing occurred during the latter half of the activity cycle (afternoon in patients and at ZT18 in mice). While OVA sensitization in mice cannot recapitulate all aspects of human asthma, the data suggest that the time-of-day variation in omalizumab effectiveness observed in patients has a biological basis. Importantly, the clinical benefit of later-day dosing was substantial: for every hour initial omalizumab treatment was delayed from 8:00 to 17:00 PM, the adjusted odds of a treatment response increased by 34%, translating to a number needed to treat (NNT) of 4 in our cohort. These results were robust across independent statistical approaches and sensitivity analyses. Interestingly, no association was observed for doses 2–3, but a time effect statistically re-emerged for dose 4 in time-to-event analyses. This may reflect the pharmacology of omalizumab, as this biologic reaches steady state after about four half-lives in patients dosed monthly (the majority of patients in our cohort).[23, 39–42] Regardless, asthma exacerbation trajectories diverged quickly for afternoon vs. morning groups based on the time of day of dose 1, becoming nearly parallel by after day 90. This reinforces the primacy of treatment initiation in dictating how time of day influences therapeutic outcomes.

Our data should be interpreted in the context of known circadian variations in asthma symptomatology, physiology, and pharmacology. Nighttime symptoms are often the first evidence of lack of asthma control, and circadian variation in autonomic function contributes to nocturnal asthma.[9, 43] Airway caliber and the central hypercapnic respiratory response oscillate cyclically with increased airway resistance and tolerance of hypercapnia in the early morning.[44, 45] In allergic asthma, airway hyperresponsiveness to inhaled triggers is magnified in the evening,[46] and some studies indicate that inhaled corticosteroids are more effective when administered in the evening.[47] Our study is notable in extending known biological rhythms in asthma to include the response to a biologic agent (omalizumab). The fact that anti-IgE therapy appears more effective when given later in the day is logical, since it might enable initial peak antibody levels to occur when asthma-related lung pathology is predicted to be at its worst. We speculate this might set off a “virtuous cycle” that yields therapeutic dividends for the subsequent course of treatment. More research is needed to understand the mechanisms underlying our observations.

Our observations are reminiscent of a growing body of literature reporting diurnal rhythms in a diverse cancer immunotherapies including immune checkpoint inhibitors, hematopoietic stem cell transplantation (HSCT), and chimeric antigen receptor (CAR) T cells.[48] For these treatments, morning dosing was associated with an improved therapeutic index, in contrast to our findings with omalizumab. We speculate this reflects mechanistic differences between omalizumab that targets mast cell activation and cancer immunotherapies that are directed at T cells.

Murine model data supported our clinical observations and revealed a multifaceted relationship between the time of day and the ability of IgE antagonism to attenuate airway inflammation. First, OVA-induced type 2 lung inflammation was itself time-of-day sensitive, with . This is consistent with studies documenting circadian clock modulation of various fundamental immune processes during vaccination.[8, 49] In a mouse model of allergic food allergy that employed OVA sensitization, OVA challenge at 17:00 local time (likely corresponding to ZT11) produced more severe diarrhea symptoms and type-2 cytokine induction than OVA challenge at 05:00 (likely ZT23), which is similar to the time effect we observed with intranasal OVA exposure.[50] Notably, circadian rhythm misalignment induced by lesioning the suprachiasmatic nucleus (SCN) eliminated time-of-day differences in the OVA food allergy model. [50] The proposed mechanism in this study involved time-of-day variation in epithelial barrier integrity, which could lead to differential translocation of the allergic trigger into the submucosal space where mast cells accumulate.[50] More recently, circadian rhythms in epithelial lung epithelial barrier integrity were observed in mouse primary airway epithelial cell culture.[51] It is worth noting that time effects in allergic inflammation are not restricted to OVA but were also reported for house dust mite exposure in mice, again with treatment at ZT12 generating the highest levels of type-2 inflammation.[52]

We found that anti-IgE dosing at ZT18 produced the most consistent attenuation of OVA-induced pathology, followed by ZT12 and ZT6, whereas anti-IgE dosing at ZT0 was ineffective. Altogether, this effectiveness gradient aligned with a similar trend in patients, accounting for the fact that humans are diurnal organisms and mice are nocturnal. However, our mouse data also emphasize that seemingly straightforward time-of-day observations in patients likely belie considerable biological complexity that may be difficult to fully anticipate or control for in real-world clinical settings.

This study has several limitations. As with any retrospective study, we cannot rule out unmeasured factors that may account for our findings. A high proportion of patients had no exacerbations in the year before baseline suggesting we might not capture all exacerbations in this cohort of patients or patients might be using stockpiles of oral steroids at home. While this did not differ by afternoon vs. morning group and we conducted robust sensitivity analyses as well as performed validation experiments in mice, our retrospective observations will need to be replicated and prospectively validated. Almost all our cohort (98%) received their doses between 8:00-17:00. As such, we are unable to extrapolate outside this time-frame. That said, our mouse model simulated anti-IgE treatment throughout the entire circadian cycle and yielded conclusions consistent with the clinical observations. We limited our cohort to patients for whom we could extract omalizumab administration times. These patients may differ from patients for whom timestamps were unavailable. However, our research question compared patients with documented dosing times, which would be less likely to be affected by these factors. Our preclinical study in mice using a single-allergen model cannot simulate all aspects of human asthma and should be validated in complementary experimental systems that examine asthma pathogenesis beyond the type-2^hi^ endotype modeled here and establish whether the circadian clock is required for our observation.

In conclusion, our findings point to a potentially simple and cost-neutral way of optimizing omalizumab, and potentially other respiratory biologics, therapy by targeting treatment to immunologically optimal times of day (chronotherapy). We suggest evaluating asthma biologics chronotherapy in pragmatic randomized controlled trials.

## DATA AVAILABILITY STATEMENT

No new data were generated for the clinical portion of this manuscript. The data used cannot be shared publicly due to HIPAA regulations as enforced by the Mass General Brigham (MGB) IRB #2023P003017. The data will be shared on collaborative request and if approved by the MGB IRB. The mouse study is governed by the Washington University School of Medicine St. Louis (WUSTL) IRB #24-0180. The data will be shared on a collaborative request and if approved by the WUSTL IRB.

## AUTHOR CONTRIBUTIONS

AA and JAH conceived and designed the study, drafted the manuscript, and supervised the study. AA, GL, JH, CG, EO, and JAH contributed to data acquisition. AA, GL, JH, CG, EO, and JAH contributed to data analysis. All authors contributed to data interpretation and critically revised the manuscript. All authors approved the final version. AA and JAH take responsibility for the cohort study and the murine study respectively.

