## Supplemental File for "Time-of-Day–Dependent Effectiveness of Anti-IgE Therapy in Asthma: Clinical and Experimental Evidence"

**Supplementary Tables and Figures**

**Figure S1**: Distribution of omalizumab dosing interval in the cohort in the first year after omalizumab prescription.

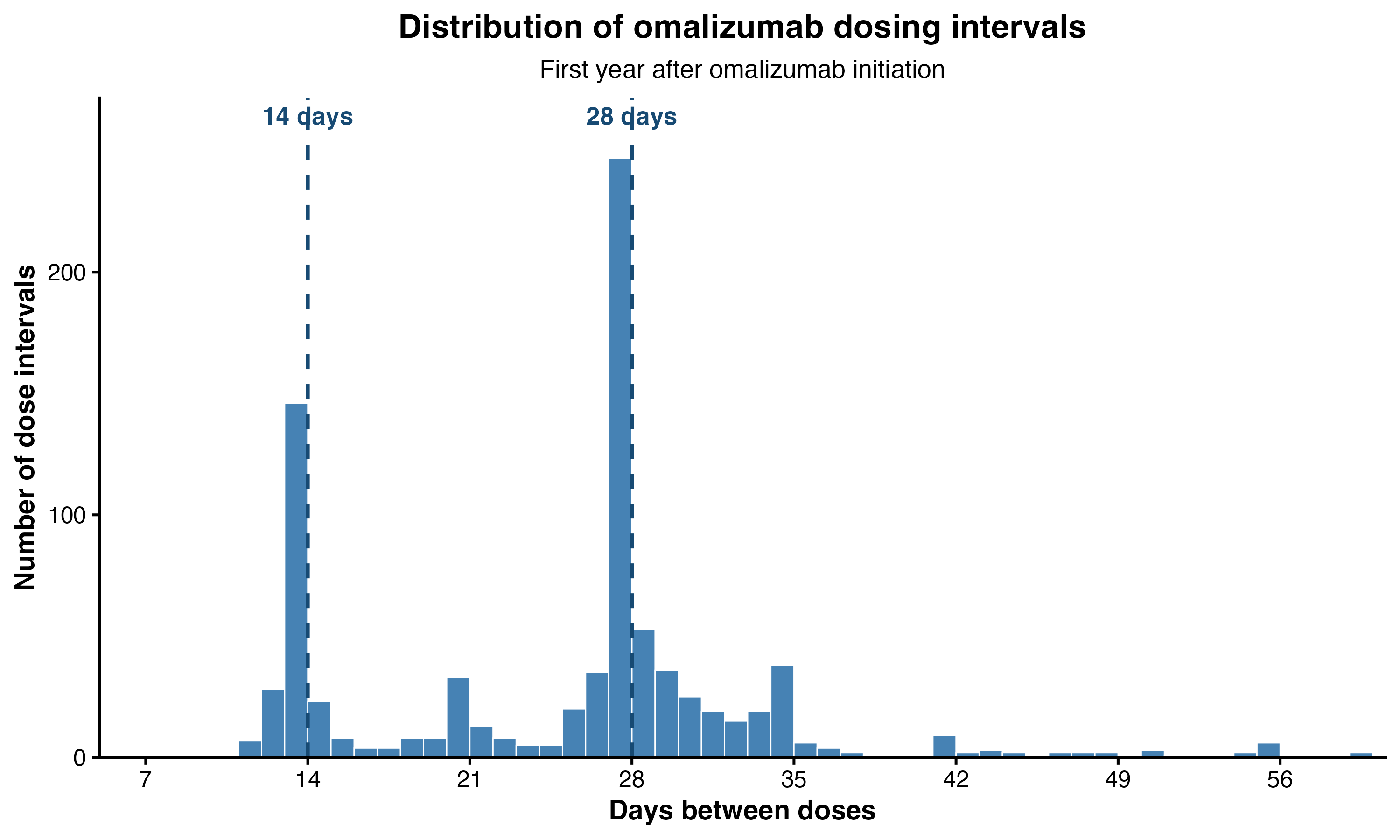

**Figure S2**: Absolute standardized mean difference before and after propensity score weighting to the overlap population.

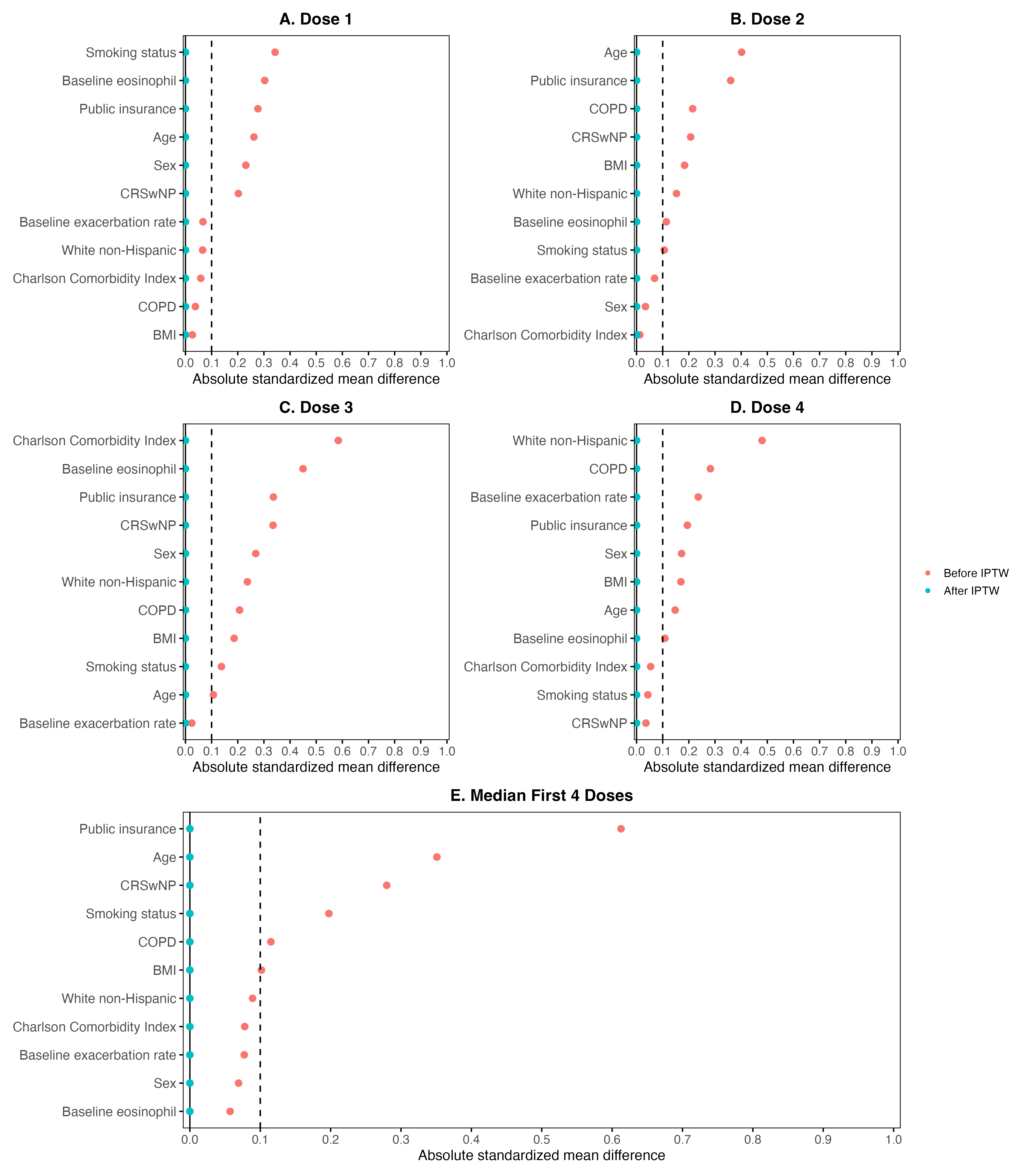

**Figure S3A,B**: (A) Predicted probability of response over 12 months based on timing of dose 1 (afternoon vs. morning) from propensity score overlap-weighted analyses and (B) Marginal probability of response based on hour of dosing.

**(3A.)**

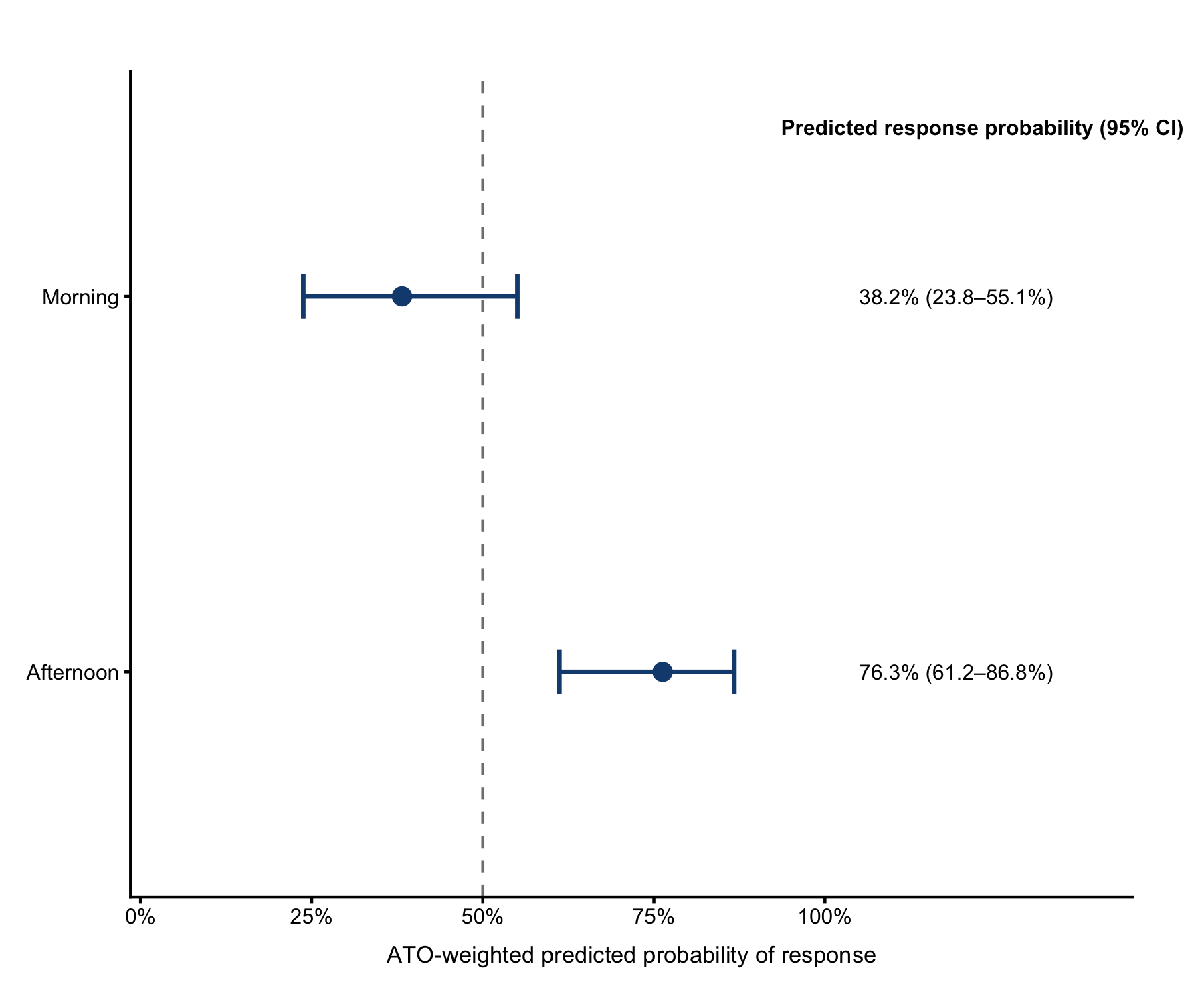

**(3B.)**

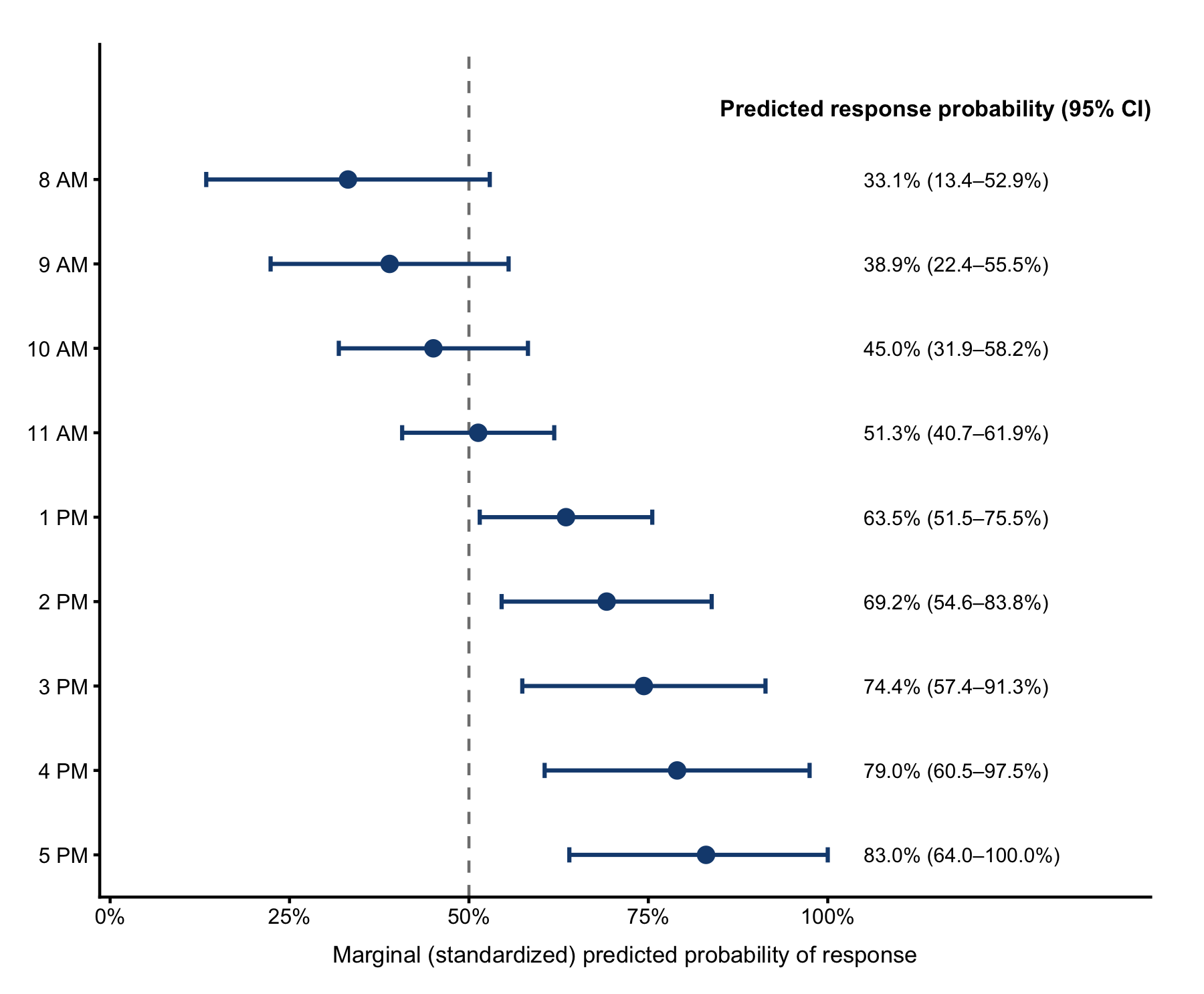

**Figure S4**: Adjusted time to first exacerbation by time of administration of (A.) Dose 2; (B.) Dose 3; (C.) Dose 4.

**B. Dose 3**

A. **Dose 2.**

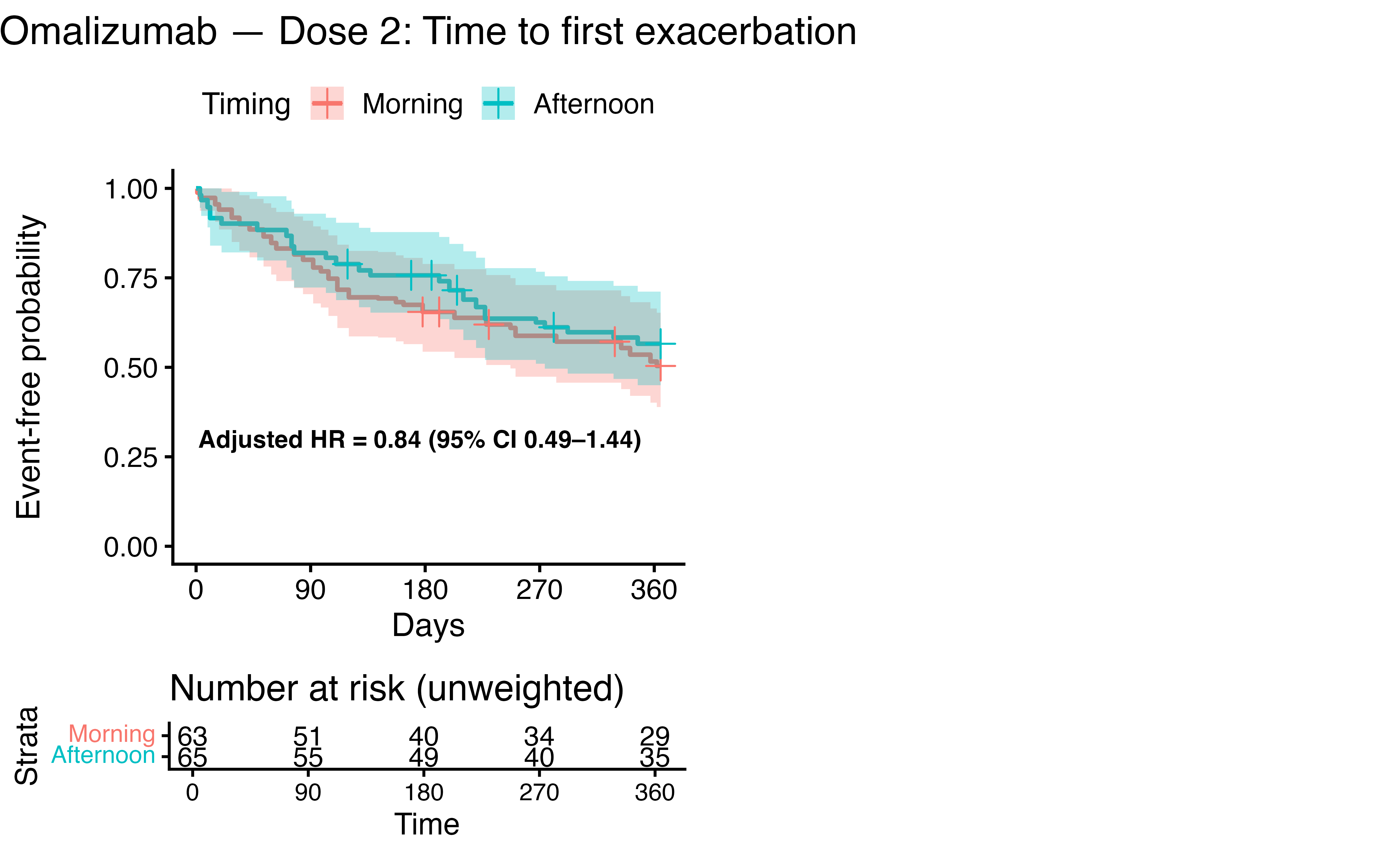

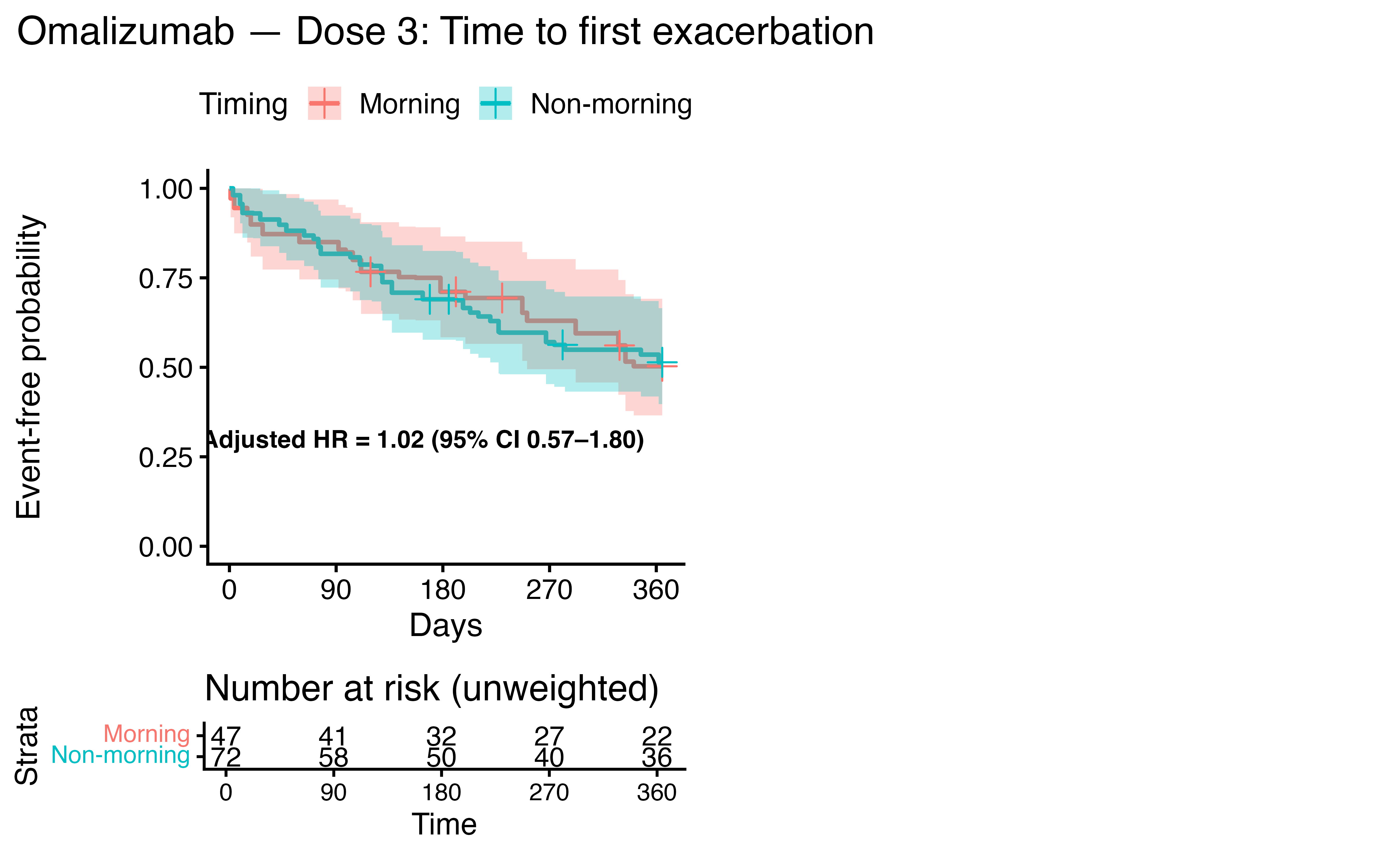

**C. Dose 4**

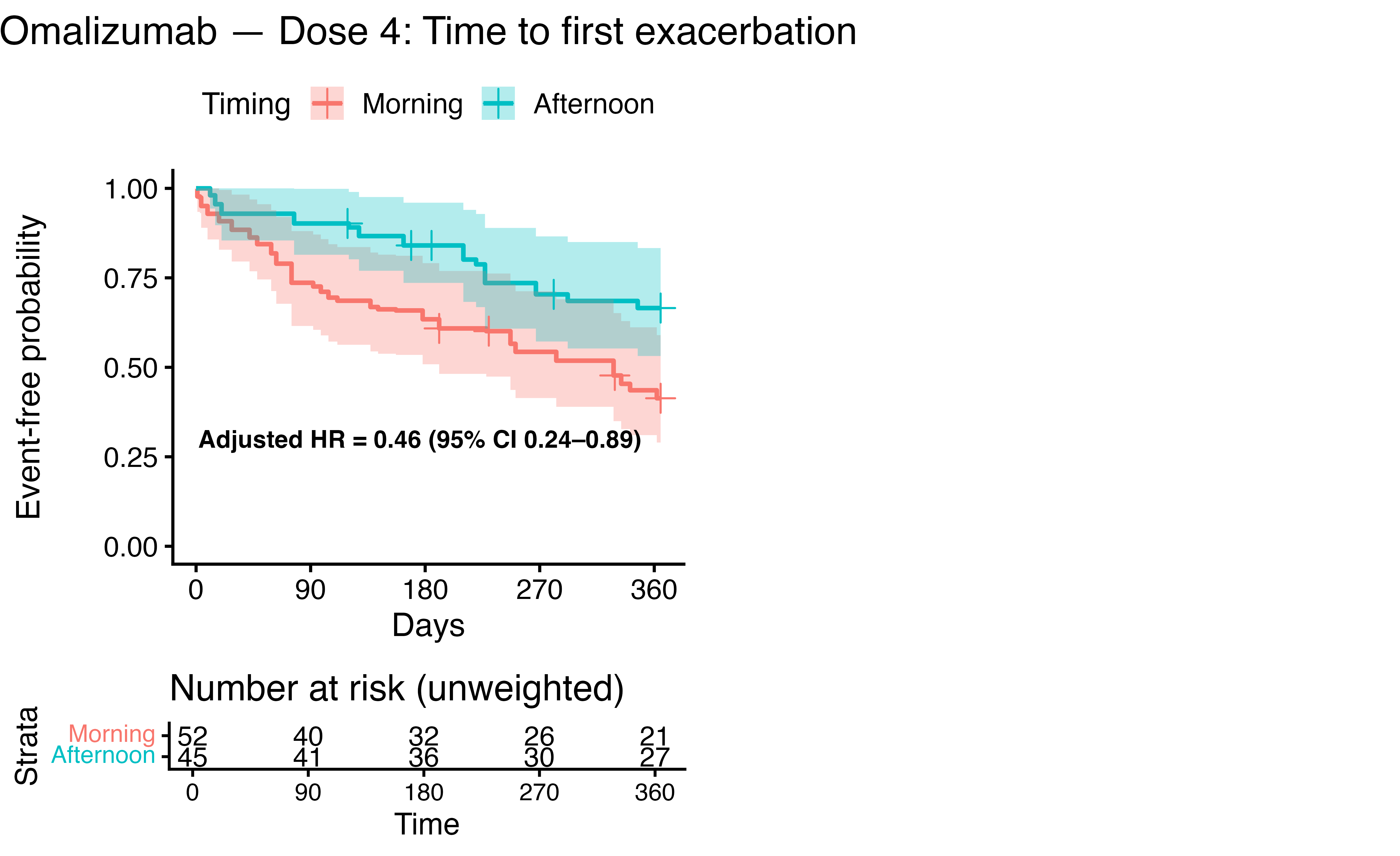

**Figure S5**: **Odds ratios for treatment response comparing afternoon vs. morning administration of dose 1, stratified by site of therapy initiation.**
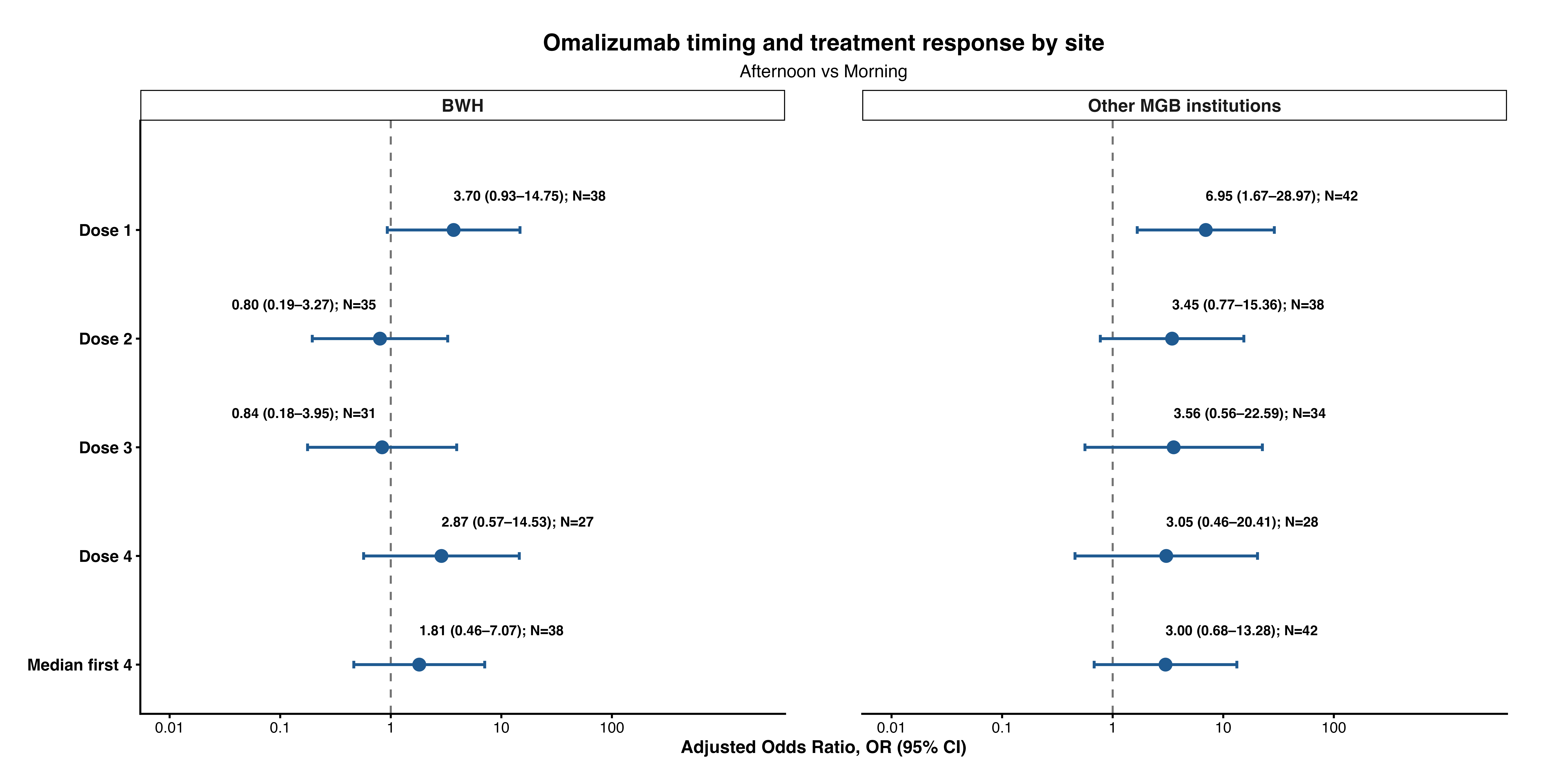

**Figure S6**: **Incidence rate ratios of exacerbations comparing afternoon vs. morning administration of dose 1, stratified by site of therapy initiation.**
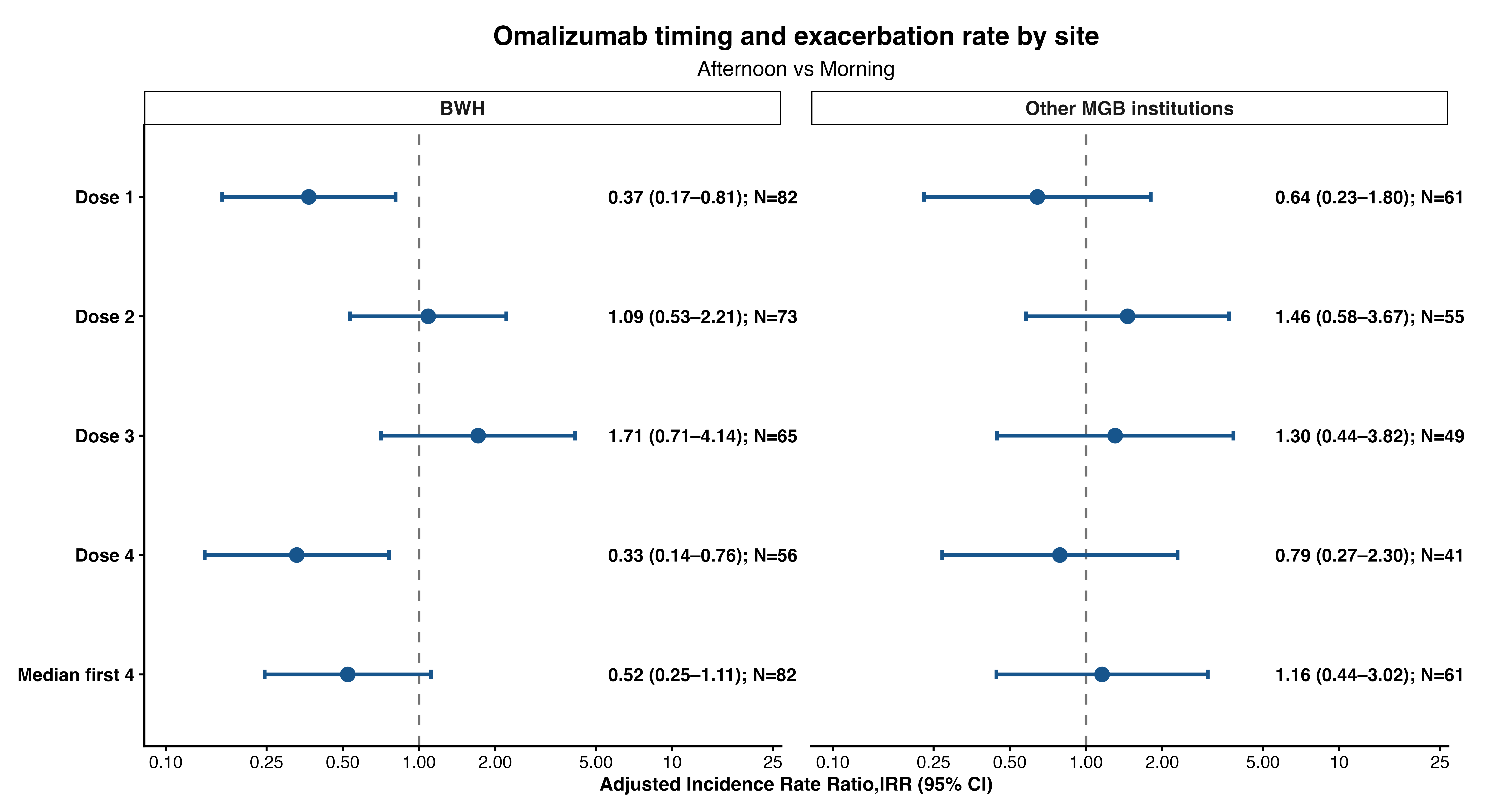

**Figure S7**: **Hazard ratios for asthma exacerbation comparing afternoon vs. morning administration of dose 1, stratified by site of therapy initiation.**

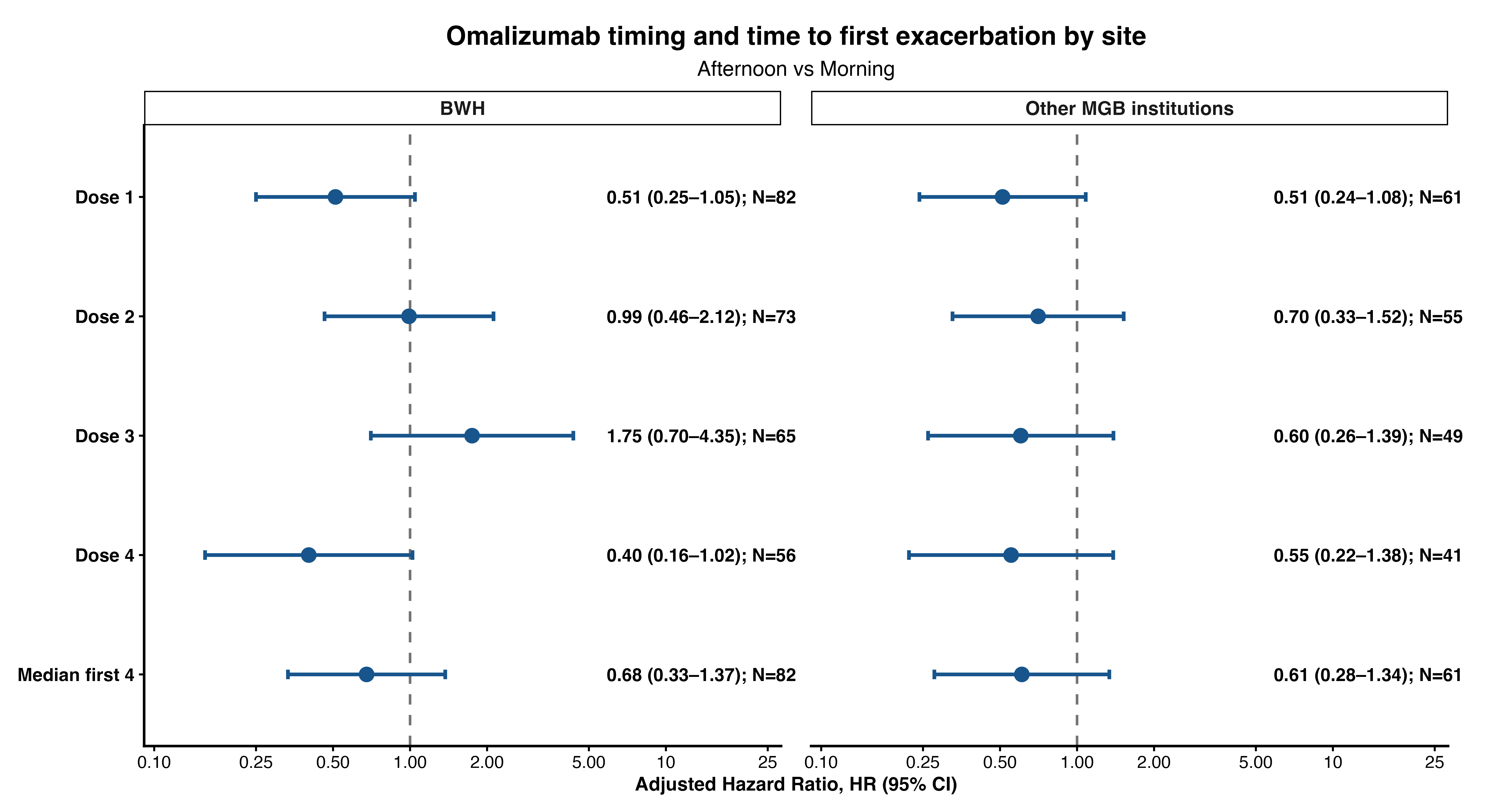

**Table S1: qPCR primers.**

| **Target** | **Vendor** | **Cat #** |
| --- | --- | --- |
| *Arg1* | IDT | Mm.PT.58.8651372 |
| *Cd4* | IDT | Mm.PT.58.12602035 |
| *Eotaxin/Ccl11* | IDT | Mm.PT.58.28587819 |
| *Ifng* | IDT | Mm.PT.58.41769240 |
| *IL-5* | IDT | Mm.PT.58.41498972 |
| *IL-6* | IDT | Mm.PT.58.10005566 |
| *IL-13* | IDT | Mm.PT.58.31366752 |
| *IL-33* | IDT | Mm.PT.58.12022572 |
| *Muc5ac* | IDT | Mm.PT.58.42279692 |
| *Tbp* | IDT | Mm.PT.39a.22214839 |

**Table S2: Unadjusted exacerbation rates and adjusted incidence rate ratios (IRR) following therapy initiation across groups**

|  | **Morning Group** | | | **Afternoon Group** | | | **Ratio of Incidence Rate Ratios (95% CI)**  **[Afternoon Group vs. Morning Group]** |
| --- | --- | --- | --- | --- | --- | --- | --- |
|  | **Baseline exacerbation rate** | **Rate over 1-year of treatment** | **IRR, post vs. pre** | **Baseline exacerbation rate** | **Rate over 1-year of treatment** | **IRR, post vs. pre** |  |
| **Dose 1** | **1.26** | 1.22 | **0.97 (0.71-1.44)** | **1.33** | 0.69 | 0.52 (0.36–0.76) | 0.52 (0.31–0.87) |
| Dose 2 | 1.47 | 0.78 | 0.53 (0.39-0.73) | 1.14 | 0.86 | 0.75 ( 0.50- 1.12) | 1.41 (0.85-2.33) |
| Dose 3 | 1.33 | 0.67 | 0.51 (0.35- 0.74) | 1.34 | 0.99 | 0.74 ( 0.51-1.06 | 1.46 (0.86-2.45) |
| Dose 4 | 1.45 | 1.06 | **0.73 (0.51-1.05)** | **1.32** | 0.60 | 0.46 ( 0.26- 0.81) | 0.63 (0.32-1.22) |

**Table S3**: **Sensitivity Analyses- Including serum IgE in the Dose 1 model.**

| Outcome | Estimate: Afternoon vs morning | Without sIgE | With sIgE | AIC before | AIC after | LRT p-value |
| --- | --- | --- | --- | --- | --- | --- |
| Binary response | Odds ratio, OR | 3.56 (1.17–10.82) | 3.61 (1.14–11.41) | 27.8 | 30.0 | 1.00 |
| Exacerbation rate | Incidence rate ratio, IRR | 0.57 (0.26–1.29) | 0.55 (0.24–1.26) | 151.5 | 153.0 | 0.48 |
| Time to first exacerbation | Hazard ratio, HR | 0.53 (0.29–0.95) | 0.54 (0.30–0.98) | 173.4 | 174.9 | 0.50 |

| Timing | Model | Measure | Estimate | LCL | UCL | P-value |
| --- | --- | --- | --- | --- | --- | --- |
| **Imputing IgE** | | | | | | |
| **Dose 1** | **Logistic** | **OR** | **5.17** | **1.88** | **14.26** | **0.001** |
| Dose 1 | NB | IRR | 0.49 | 0.24 | 1.01 | 0.053 |
| **Dose 1** | **Cox** | **HR** | **0.51** | **0.30** | **0.87** | **0.013** |
| Dose 2 | Logistic | OR | 1.50 | 0.55 | 4.11 | 0.428 |
| Dose 2 | NB | IRR | 1.26 | 0.68 | 2.34 | 0.459 |
| Dose 2 | Cox | HR | 0.84 | 0.49 | 1.44 | 0.524 |
| Dose 3 | Logistic | OR | 1.40 | 0.44 | 4.41 | 0.566 |
| Dose 3 | NB | IRR | 1.51 | 0.78 | 2.92 | 0.218 |
| Dose 3 | Cox | HR | 1.03 | 0.57 | 1.88 | 0.913 |
| Dose 4 | Logistic | OR | 2.41 | 0.71 | 8.21 | 0.159 |
| Dose 4 | NB | IRR | 0.54 | 0.25 | 1.14 | 0.104 |
| **Dose 4** | **Cox** | **HR** | **0.46** | **0.24** | **0.90** | **0.022** |
| Median first 4 | Logistic | OR | 2.18 | 0.79 | 6.01 | 0.133 |
| Median first 4 | NB | IRR | 0.75 | 0.38 | 1.49 | 0.415 |
| Median first 4 | Cox | HR | 0.64 | 0.38 | 1.08 | 0.097 |
| **Imputing IgE and Eosinophil count** | | | | | | |
| **Dose 1** | **Logistic** | **OR** | **4.36** | **1.64** | **11.61** | **0.003** |
| Dose 1 | NB | IRR | 0.51 | 0.26 | 1.01 | 0.053 |
| **Dose 1** | **Cox** | **HR** | **0.59** | **0.35** | **0.98** | **0.042** |
| Dose 2 | Logistic | OR | 1.46 | 0.54 | 3.89 | 0.455 |
| Dose 2 | NB | IRR | 1.27 | 0.69 | 2.34 | 0.440 |
| Dose 2 | Cox | HR | 0.86 | 0.50 | 1.48 | 0.589 |
| Dose 3 | Logistic | OR | 1.22 | 0.39 | 3.82 | 0.730 |
| Dose 3 | NB | IRR | 1.59 | 0.83 | 3.05 | 0.163 |
| Dose 3 | Cox | HR | 1.12 | 0.61 | 2.04 | 0.719 |
| Dose 4 | Logistic | OR | 2.24 | 0.70 | 7.15 | 0.175 |
| Dose 4 | NB | IRR | 0.54 | 0.26 | 1.10 | 0.087 |
| **Dose 4** | **Cox** | **HR** | **0.48** | **0.25** | **0.92** | **0.027** |
| Median first 4 | Logistic | OR | 1.95 | 0.72 | 5.26 | 0.189 |
| Median first 4 | NB | IRR | 0.79 | 0.41 | 1.53 | 0.486 |
| Median first 4 | Cox | HR | 0.72 | 0.43 | 1.21 | 0.220 |

**Table S4: Sensitivity Analyses- Imputing IgE and Eosinophil count for individuals in which they were missing using multiple imputation by chained equations (mice).**

**Table S5: Sensitivity Analyses- Including Season (Spring, Summer, Winter, Fall) in the Dose 1 model**

| Outcome | Effect estimate: Afternoon vs morning | Without season | With season | AIC without season | AIC with season | LRT p-value |
| --- | --- | --- | --- | --- | --- | --- |
| Binary response | Odds ratio, OR | 5.20 (1.94-13.96) | 5.30 (1.95-14.40) | 37.3 | 40.5 | 0.43 |
| Exacerbation rate | Incidence rate ratio, IRR | 0.49 (0.24-0.99) | 0.48 (0.26-0.89) | 188.3 | 192.4 | 0.60 |
| Time to first exacerbation | Hazard ratio, HR | 0.52 (0.31–0.87) | 0.53 (0.31–0.89) | 230.4 | 235.2 | 0.75 |

**Table S6: Subgroup Analyses of Treatment Response Comparing Afternoon vs. Morning Administration of Dose 1, Stratified by Site of Therapy Initiation**

| Outcome | BWH (n= 87)  Estimate (95% CI) | Other MGB hospitals (n=66)  Estimate (95% CI) | P-value for interaction |
| --- | --- | --- | --- |
| **Response** | **OR 3.70 (0.93- 14.7)** | **OR 6.95 (1.67- 29.0)** | **0.55** |
| **Time to exacerbation** | **HR 0.51 (0.25- 1.05)** | **HR 0.51 (0.24-1.08)** | **1.00** |
| **Exacerbation rate** | **IRR 0.37 (0.17- 0.81)** | **IRR 0.64 (0.23- 1.80)** | **0.40** |

**Table S7**: **Sensitivity Analyses- Including Splines and Circular Time in the Dose 1 model.**

| **Timing exposure** | **Spline vs Linear ΔAIC** | **Spline LRT P-value** | **Circular vs linear ΔAIC** |
| --- | --- | --- | --- |
| Dose 1 | 0.63 | 0.185 | 0.54 |
| Dose 2 | 3.58 | 0.811 | 2.05 |
| Dose 3 | 0.66 | 0.188 | 2.12 |
| Dose 4 | 3.89 | 0.947 | 2.05 |
| Median first 4 doses | 0.84 | 0.206 | 2.36 |
| *Δ= change in; AIC, Akaike Information Criterion; LRT, Likelihood Ratio Test | | | |

**Additional Methods.**

1. **Retrospective cohort study**

**1.1. Stratified and Sensitivity Analyses.** We evaluated if our results were consistent when stratified by site of administration across the main hospitals within the MGB health system (Brigham and Women’s Hospital, which houses the Severe Asthma Program, vs. other hospitals) and by dosing interval (biweekly or monthly), classifying patients were classified according to their dominant dosing schedule during the first year of therapy. To do this, we classified the interval between every pair of consecutive doses a patient received into biweekly (if the interval was <21 days) and monthly if ≥ 21 days. We excluded doses where the interval was less than five days or more than 60 days. Thereafter, we classified patients based on the mode of the intervals between their doses. Two patients with equal numbers of biweekly and monthly-classified intervals between their doses were adjudicated as biweekly based on their observed dosing patterns (#1: intervals of 14 and 22; #2: intervals of 21, 21, 22, 23, and 68). Patients with only one recorded administration (n=16), for whom a dosing interval could not be determined, were also excluded from the dosing-interval subgroup analyses. We first evaluated effect modification by dosing schedule using an interaction term rather than fitting separate models for the dosing subgroups. However, estimates were unstable when adjusted. Thus, we restricted the analyses to patients with a predominantly monthly dosing schedule (n=104), as the small number of patients with predominantly biweekly dosing (n=33) led to unstable weighted estimates.

**1.2. Missing Data and Multiple Imputation**. Missing blood eosinophil values were addressed using We used multiple imputation by chained equations (MICE) under the missing-at-random assumption to impute missing blood eosinophil levels and immunoglobulin E (IgE) levels. Of 153 patients, 10 (6.5%) had missing eosinophil measurements, 28 (18.3%) had missing IgE, and 2 (1.3%) missing both IgE and eosinophil measurements. We generated 50 imputed datasets using predictive mean matching, with 20 iterations per imputation. The imputation models included omalizumab administration time, covariates as included in the primary regression models, which include demographic characteristics, baseline exacerbation history, and comorbid conditions, and observed exacerbation outcomes given that these missing features may be related to follow-up outcomes. Analyses were repeated across the imputed datasets and estimates were pooled using Rubin's rules to account for within- and between-imputation variability.

2. **Murine study**

**Mice.** Six-week-old female BALB/C mice were obtained from Jackson Laboratory and housed in cages with food available *ad libitum*. In circadian rhythm research, internal biological time is represented in “Zeitgeber Time” (ZT) units, where ZT0 is lights-on, and ZT12 is lights-off.[32] The mice were first habituated to one of four staggered light-dark cycles (ZT0, ZT6, ZT12, ZT18) before OVA sensitization to adjust their circadian phases.[33] Mouse actigraphy was recorded in using passive IR monitors and data were analyzed using ClockLab software (Actimetrics).

**OVA sensitization.** We employed an adjuvant-free OVA sensitization protocol to experimentally model allergic airway inflammation (**Fig 2, A**).[34] OVA sensitization commenced after mice acclimatized for two weeks in custom light cycles to simulate different exposure times of day. All treatments were administered at 12:00 local time under dim red lighting to avoid blue-light exposure that could shift the mice's circadian phase. Mice were anesthetized with 2% isoflurane and received subcutaneous injections of OVA (10 mg/100 mL sterile PBS) on days 0, 7, and 14. On Days 26, 27, and 28, they were anesthetized and intranasally dosed with 20 mg OVA in 30 mL of sterile PBS. Mice received a single dose of anti-IgE monoclonal antibody at ZT0, ZT6, ZT12, or ZT18, just before the first OVA inhalational exposure, thereby simulating dose 1 of omalizumab. To simulate omalizumab therapy, mice received a single (100 mg/100 mL) dose of anti-IgE antibody (clone R35-72, BD Biosciences). Sham (PBS) injected mice served as controls.

**Respiratory system mechanics** were measured using a Buxco Elan RC mouse volume-controlled ventilator as described.[35] Briefly, mice were anesthetized with an intraperitoneal (i.p.) injection of a ketamine/xylazine cocktail, and then a tracheostomy was performed using a 20G flexible catheter. Mice were then connected to the ventilator, and 30 mL of PBS was administered via an in-line nebulizer, followed by methacholine at escalating concentrations (Sigma-Aldrich). Dynamic respiratory system inspiratory resistance (R_I_) was then measured over a 5-minute epoch, and the average value was recorded.

**Quantitative PCR (qPCR).** RNA extraction and qPCR were performed using standard protocols.[35] Lungs were manually excised and immersed in RNAlater solution (Applied Biosystems). Total RNA was isolated from lung homogenates using RNeasy Mini Kits (Qiagen) or TRIzol (Thermo Fisher). Reverse transcription was carried out using the Applied Biosystems High-Capacity cDNA Reverse Transcription Kit. Real-time qPCR was performed on the StudioQuant 5 System (Applied Biosystems). Primers used for qPCR are listed in **Table S1**.

**Histology and Immunostaining.** Lungs were fixed by inflation with 10% buffered formalin at 20 cmH_2_O, paraffin-embedded, and stained with Periodic Acid Schiff Stain.[36] Slides were scanned using a Hamamatsu Nanozoomer at the Alafi Neuroimaging lab at Washington University. PAS-positive and hematoxylin-positive pixels were counted using Aperio ScanScope Software.[37]
